# Persisting Salivary IgG against SARS-CoV-2 at 9 Months After Mild COVID-19: A Complementary Approach to Population Surveys

**DOI:** 10.1101/2021.03.13.21253492

**Authors:** Hassan Alkharaan, Shaghayegh Bayati, Cecilia Hellström, Annika Olsson, Karin Lindahl, Gordana Bogdanovic, Soo Aleman, Georgios Tsilingaridis, Patricia De Palma, Sophia Hober, Anna Månberg, Peter Nilsson, Elisa Pin, Margaret Sällberg Chen

**Affiliations:** Karolinska Institutet, Department of Dental Medicine, 14152, Stockholm, Sweden; Karolinska University Hospital, Department of Infectious Diseases, 14186, Stockholm, Sweden; Karolinska University Hospital, Department of Clinical Microbiology, 14186, Stockholm, Sweden; Karolinska Institutet, Department of Medicine, 14186, Stockholm, Sweden; Division of Affinity Proteomics, Department of Protein Science, School of Engineering Sciences in Chemistry, Biotechnology and Health (CBH), KTH Royal Institute of Technology, SciLifeLab, Stockholm, Sweden; Division of Protein Technology, Department of Protein Science, KTH Royal Institute of Technology, Stockholm, Sweden

**Keywords:** COVID-19, saliva, antibody, serology, convalescence, immunoassay

## Abstract

**Background:** Declining humoral immunity in COVID-19 patients and possibility of reinfections has raised concern. Mucosal immunity particularly salivary antibodies could be short-lived. However, long-term studies are sparse.

**Methods:** Using a multiplex bead-based array platform, we investigated antibodies specific to severe acute respiratory syndrome coronavirus 2 (SARS-CoV-2) proteins in 256 saliva samples from convalescent patients 1-9 months after symptomatic COVID-19 (n=74, Cohort 1), undiagnosed individuals with self-reported questionnaires (n=147, Cohort 2), and individuals sampled pre-pandemic time (n= 35, Cohort 3).

**Results:** Salivary IgG antibody responses in Cohort 1 (mainly mild COVID-19) were detectable up to 9 month recovery, with high correlations between spike and nucleocapsid specificity. At 9 months, IgG remained in saliva in majority as seen in blood serology. Salivary IgA was rarely detected at this timepoint. In Cohort 2, salivary IgG and IgA responses were significantly associated with recent history of COVID-19 like symptoms. Salivary IgG also tolerated temperature and detergent pre-treatments.

**Conclusions:** Unlike SARS-CoV-2 salivary IgA that appeared short-lived, the specific IgG in saliva appears stable even after mild COVID-19 as noted for blood serology. The non-invasive saliva-based SARS-Cov-2 antibody testing with self-collection at homes may thus serve as a complementary alternative to conventional blood serology.

## Introduction

The novel severe acute respiratory syndrome coronavirus 2 (SARS-CoV-2) outbroke in an abrupt fashion since it was reported in Wuhan, China, in late December 2019 (1), and obligated the World Health Organization to declare a global health emergency which escalated the concern to a pandemic situation on March 2020. As of Mars 2021, SARS-CoV-2 had infected over 114 million cases and caused up to 2.5 million global deaths (2). The human adaptive immune system plays a key role to eliminate and memorize infectious microbes by launching a cascade of physiological activities that brings to the activation of B and T lymphocytes. B lymphocytes are responsible for producing antibodies that recognize and neutralize SARS-CoV-2 antigens in order to eradicate the infection and have shown to play a vital role in protecting against re-infections in animal and humans (3–5). IgG, IgA, and IgM antibodies, all principal contributors of humoral immunity, are activated against SARS-CoV-2 and detected in the circulating blood of more than 90% of infected individuals from the 11-13 day post-symptom onset (PSO) (6–8). Currently, the SARS-CoV-2 immunity is under extensive examinations; a recent study showed that circulating antibodies post-SARS-CoV-2 infection can persist up to 8 months (9), and other previous studies show this immunological memory persists for a certain period followed by a slight decay, especially in asymptomatic infected individuals (10–14). Oral and nasal cavities are considered the main gate for SARS-CoV-2 virus entry, and saliva secretory antibodies may be the first immunity arm that combat the infection by recognizing the virus. Salivary antibodies to SARS-CoV-2 can be detected early after symptom onset and may persist for up to at least three months post infection (8,10,12). Hence, saliva sampling could be a sensible and non-invasive way to indicate SARS-CoV-2 exposure. Similar to the previous SARS-CoV and MERS-CoV, the spike protein (S) of SARS-CoV-2 recognizes the angiotensin-converting enzyme 2 (ACE2) receptor and uses it as a key entry to attack the host cells (15–17). Antibodies play an important role in resolving acute SARS-CoV-2 infection (11,18) but differential features of anti-SARS-CoV-2 antibodies negatively impacting COVID-19 severity in hospitalized patients is also described, especially related to complement deposition and systemic inflammation (19). Understanding the dynamics and durability of antibody memory to SARS-CoV-2 is an instrumental step to manage the pandemic, and even useful in deploying vaccination strategies. However, as the mucosal immunity is known to be short-lived, the durability of SARS-CoV-2 specific antibodies in saliva could be limited, and whether they permit detection 3-4 month after infection (8,10) is of great interest.

In this study, we exploited a highly sensitive and specific multiplex SARS-CoV-2 serology platform previously validated for seroprevalence studies (20) to investigate SARS-CoV-2 antibodies in the saliva. Samples from patients with diagnosis of mild coronavirus disease (COVID-19) in the convalescence phase, at 1-9 months after diagnosis of COVID-19; and from undiagnosed individuals with or without history of COVID-19 symptoms, were analysed and compared to pre-pandemic samples. Our data indicate that spike-specific IgG reactivity is detectable in saliva in vast majority of patients 1 - 9 months post infection. This result was similar to that detected by blood serology performed in the clinical diagnostic laboratory. The IgA reactivity on the other hand was short-lived in saliva, detectable only the first 3 months. Moreover, IgG and IgA reactivity to both spike and nucleocapsid antigens significantly correlated with a history of COVID-19 like symptoms in undiagnosed individuals.

## Materials and methods

### Experimental Design

We applied a bead-based serology assay to detect IgG and IgA to SARS-CoV-2 proteins in saliva samples to evaluate its performance. The assay method is originally developed for detection of SARS-CoV-2 specific IgG in serum and plasma (20). Salivary IgG and IgA responses to five different SARS-CoV-2 antigens (three spike proteins and two nucleocapsid proteins) were first tested. The antigens’ performance in classifying positive and negative samples was evaluated for the single antigens as well as for antigens combined in panels. Best performing representations of spike and nucleocapsid were chosen in subsequent assessments.

### Cohort Design

The study was approved by the human ethical authority (dnr 2020-01702, 2020-06381) and complied with the declaration of Helsinki. All participants were recruited after signing informed consent forms for this observational study. Saliva samples (total n=256) were collected and arranged in following groups. Cohort 1: convalescence COVID-19 samples (n=74) of 72 patients (2 participants donated twice at 6 months apart) diagnosed with COVID-19 during March-April 2020, were collected from June to December 2020; Cohort 2: samples from undiagnosed individuals donated during May-Nov 2020 (n=147); Cohort 3: anonymous saliva samples were from 2018 before the COVID-19 outbreak (pre-pandemic, n=35).

All convalescent patients (Cohort 1) had COVID-19 diagnosis confirmed by SARS-CoV-2 RT-PCR, except one patient who had positive SARS-CoV-2 antibodies at four time points in the convalescence phase. Seroconversion was tested by clinical SARS-CoV-2 blood serology assays (described below). The patients were recruited from the department of Infectious Diseases, Karolinska University Hospital (n=65), and University Dental Clinic of Karolinska Institutet (n=7). Clinical demographic data of convalescent patients was compiled from medical journal records or questionnaire, and used in subgroup analysis. Among 72 patients, 95.8% had mild COVID-19, without hospitalization due to COVID-19 symptoms. Three were admitted to hospital for purpose of only isolation, and three were admitted due to COVID-19 symptoms. In the latter group, two were hospitalized without any required oxygen treatment and one received maximum 1.5 litre oxygen treatment during the hospitalization, indicating no severe disease outcome. The time-points of serum and saliva samples collection were grouped according to time post symptom onset (PSO), i.e. (i) PSO less than 3 months. (ii) PSO of 3-8 months. (iii) PSO of 9 months. Cohort 2 constitutes of anonymous participants visiting the premises of University Dental Clinic of Karolinska Institutet or Eastman Institute Stockholm during the study time, such as patients, staff, or relatives to them. A questionnaire was used to collect COVID-19 related symptom information for sub-group analysis of undiagnosed samples into (i) Symptomatic (ii) Non-symptomatic, based on their past 3 months health condition before sampling.

### Saliva samples collection

Expectorated unstimulated whole saliva samples were used throughout this study. All samples were self-collected using standardized instructions and sample tubes provided by this study, processed and stored at −80 within 24 hr. Salivary stability tests were performed on samples subgroups to evaluate the antibody reactivity using samples treated with 1% Triton X-100 for 1 hour at room temperature or heat-treated at 56 C for 30 min in water bath to allow viral inactivation (19). Eighteen antibody-positive from Cohort 1 and 10 antibody-negative samples from Cohort 2 were included in the comparison. Incubation at room temperature for one to three days was also tested in five samples to simulate the standard circumstances of mailed-in saliva self-collection procedure. Saliva samples from convalescent patients (Cohort 1) were collected on the same day as venous blood during a COVID-19 follow-up examination at the department of Infectious Diseases, Karolinska University Hospital.

### Clinical serology tests

Paired serum samples of all convalescent patients were tested by Dept. of Karolinska University Hospital Clinical Microbiology Laboratory. Three automated and one in-house diagnostic methods were used under the study period of included convalescence blood samples - SARS-CoV2-IgG test iFlash 1800 YHLO (CLIA), LIAISON® SARS-CoV-2 S1/S2 IgG test DiaSorin (CLIA), and SARS CoV-2 IgG in-house ELISA for samples taken prior to June 2020 mainly early convalescence samples (<9month). The Elecsys® Anti-SARS-CoV-2 antibody test Roche (ECLIA) was used for all late convalescence samples (9-month). YHLO determines the antibodies against SARS-CoV-2 nucleocapsid and spike protein, DiaSorin against spike protein, whilst Elecsys®, and in-house ELISA to the recombinant nucleocapsid protein. The tests use different techniques such as chemiluminescence immunoassay (CLIA), electrochemiluminescence immunoassay (ECLIA), and enzyme-linked immunoassay (ELISA).

### Antigen Production

The proteins were produced as following: 1) Spike-f as spike trimers comprises the prefusion-stabilized spike glycoprotein ectodomain is expressed in HEK cells and purified using a C-terminal Strep II tag, 2) Spike S1 domain was expressed in CHO cells and purified using C-terminal HPC4-tag, 3) Spike RBD domain was expressed in HEK cells and purified using the mFc C tag; 4) nucleocapsid, one full-length version and 5) one nucleocapsid C-terminal chain were each expressed in *E*.*coli* and purified using a C-terminal His-tag (21,22).

### SARS-CoV-2 antibody detection by a bead-based assay

The analysis of salivary antibodies was performed as previously described (20) with a few modifications. Briefly, each antigen was diluted to a final concentration of 80 µg/ml (100mM) with 2-(N-morpholino) ethanesulfonic acid buffer, pH 4.5 (SigmaAldrich) and immobilized on uniquely color-coded bead type (bead ID) (MagPlex-C, Luminex corp.). The antigen-immobilized beads were then pooled to form the bead array. Besides the viral antigens, anti-human IgG (309-005-082, Jackson Immunoresearch), anti-human IgA (800-338-9579, Bethyl), and the EBV EBNA1 protein (ab138345, Abcam) were also included as sample loading controls.

Saliva samples were diluted 1/5 in assay buffer composed of 3% bovine serum albumin (w/v), 5% non-fat milk (w/v) in 1×PBS supplemented with 0.05 % (v/v) Tween20 (VWR, 437082Q) and incubated with the bead array for 1 hour at room temperature and 650 RPM rotation. Afterwards, the antigen-antibody complexes were cross-linked by adding 0.2% paraformaldehyde (AlfaAesar, 30525-89-4) in PBS 0.05% Tween 20 (PBS-T) for 10 min at room temperature. Detection was performed by applying R-phycoerithryne-conjugated anti-human IgG (H10104, Invitrogen) diluted 0.4 µg/mL, or R-phycoerithryne-conjugated anti-human IgA (800-338-9579, Bethyl) diluted 0.2 µg/mL in PBS-T for 30 minutes at room temperature. Finally, the read-out was performed by using a FlexMap3D system and the xPONENT software (Luminex Corp.).

### Statistical analysis

Statistics and visualizations of the multiplex bead array generated data were performed using R (version 3.6.1) with RStudio (version1.2.1335) and the additional packages heatmap (1.0.10), reshape2 (1.4.3). In-house developed functions were used for instrument file import and quality control. The bead array results were acquired as Median Fluorescent Intensity (MFI) per sample and bead identity. A cutoff for seropositivity was calculated per antigen as the mean + 7x SD of 12 negative pre-pandemic reference samples carefully selected based on their signal intensity distribution. GraphPad Prism Version 9.0.0 (86) was used for the nonparametric comparisons Mann-Whitney test and Spearman correlation analysis. Datasets also initially underwent normality distribution testing. N1 Chi-squared test was used for comparisons of binomial datasets in MedCal software calculator. Two-sided p-values <0.05 were considered significant. Descriptive analyses were made on clinical characteristics and the number of observations, presented as numbers and percentages.

## Results

### Salivary antibody reactivity to SARS-CoV-2 proteins

The assay performance was validated by comparing the capability of each of the five antigens included in the array to classify convalescent samples <3 mo–9 mo (Cohort 1, n=74) and pre-pandemic samples (Cohort 3, n=35), of which 12 samples from Cohort 3 were used to set the assay cut-offs. Among the five antigens included in the assay, spike foldon (Spike-f) and C-terminal fragment (NC-C) showed the best performance (88% and 61% sensitivity respectively, and 100% specificity) in classifying SARS-CoV-2 saliva samples of the convalescence cohort from the pre-pandemic cohort (**Table 1**). We also evaluated the antigen panel in all possible combinations of 2 and 3 proteins, considering as *positive* a sample that showed reactivity to both antigens in a panel-of-two antigens, and at least to two out of three antigens in a panel-of-three antigens (**Table S1 and 2**). There, the best performance among the panels was reached by the Spike-f, S1, RBD triple combination, showing 70.3% sensitivity and 100% specificity. Hence, the absolute best performance was shown to be reached by Spike-f as single antigen (88% sensitivity, 100% specificity) in this COVID-19 convalescent and pre-pandemic saliva collection. IgA reactivities to the included proteins were identified only in a minority of cases, with higher prevalence of reactivity to Spike-f (12%) (**Table S3**).

**Table 1.** Single antigen specificity and sensitivity in detecting SARS-CoV-2 IgG in nvalescent saliva (1-9 month PSO) and pre-pandemic saliva samples.

|  |  | Convalescent (N=74) |  |  | Pre-pandemic (N=23)<sup>#</sup> |  |  |
| --- | --- | --- | --- | --- | --- | --- | --- |
| Antigen | Host | Sensitivity [%] | Pos | Neg | Specificity [%] | Pos | Neg |
| Spike-f | HEK | 88 | 65 | 9 | 100 | 0 | 23 |
| S1 | CHO | 61 | 45 | 29 | 100 | 0 | 23 |
| RBD | HEK | 69 | 51 | 23 | 96 | 1 | 22 |
| NC | *E. coli* | 49 | 36 | 38 | 100 | 0 | 23 |
| NC-C | *E. coli* | 61 | 45 | 29 | 100 | 0 | 23 |

<sup>#</sup>Another 12 independent pre-pandemic saliva samples were used to establish assay cut-offs.

### Serum and salivary antibody reactivity overtime post Covid-19

As shown in **Table 2**, Cohort 1 were mainly patients who have had mild COVID-19 and were grouped according to duration past their diagnosis. Some were hospitalized for isolation mainly, but none received oxygen treatment or required ventilation related treatment. All individuals were free from respiratory symptoms at the 9-month follow-up but in a minority across all three groups various general residual symptoms were still noted (data not shown). As shown in **Table 3**, the vast majority of serum samples up to 9 months convalescence were still tested positive in clinical SARS-CoV-2 serology, with high seroprevalence across the whole time span of collection. Interestingly, paired saliva samples from Cohort 1 patients tested with the multiplex bead-array showed that the positivity rate of anti-Spike-f IgG in saliva remained remarkably high and in similar range (100%-87.5%) as noted for serum antibodies (88.9%-96.9) throughout from early (<3 months) to late convalescence group (9 months) (**Table 3, Figure 1a**). However, the NC-C specific IgG in saliva dropped significantly after 3 months (from 88.9% to 60.6-50.0%). As stated earlier, specific IgA responses to these antigens were detected only in a minority of the saliva samples, and was enriched in early convalescence (<3months, 44.4% for Spike-f and 11% for NC-C), while dropping to less than 10% in later convalescent samples (p<0.01).

**Table 2.**
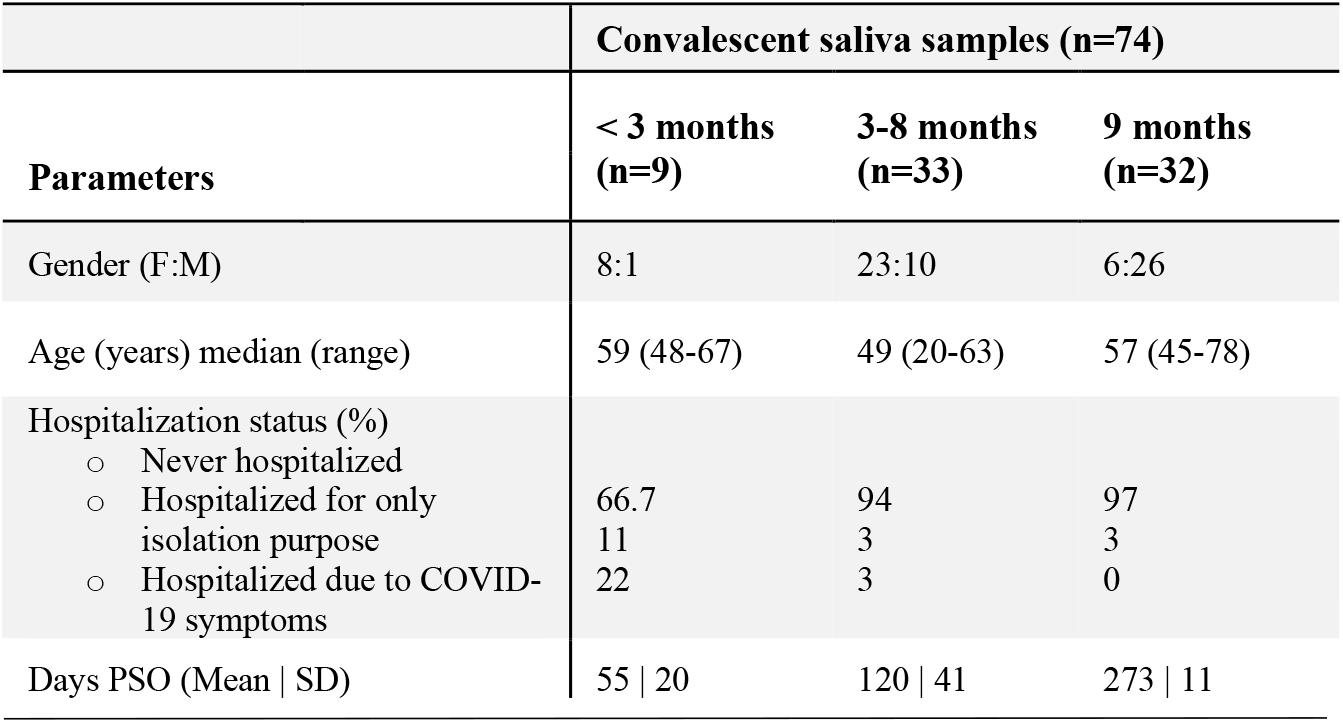
Demographic characteristics of convalescence samples of Cohort 1, grouped the time-points of post symptom onset (PSO) at which the samples were taken.

**Table 3.** Cohort 1: Salivary antibodies to Spike-f or NC-C over time concurs with rum positivity in clinically validated SARS-Cov-2 antibody diagnostics.

| Convalescence (mo) | Serum Ab SARS CoV-2 | Saliva IgG |  | Saliva IgA |  |
| --- | --- | --- | --- | --- | --- |
|  |  | Spike-f | NC-C | Spike-f | NC-C |
| <3 | 88.9% | 100.0% | 88.9% | 44.4% | 11.1% |
| 3-8 | 90.9% | 84.8% | 60.6%** | 6.3%*** | 3.1%** |
| 9 | 96.9% | 87.5% | 50.0%*** | 9.7%*** | 6.5%*** |

Note: \*\* and \*\*\* indicate $p < 0.01$ and $p < 0.0001$ respective compared to clinical SARS-Cov-2 serum antibody diagnosis, determined by N-1 Chi-squared test (Campbell I, Statistics in medicine, 2007, Richardson JTE, Statistics in medicine, 2011). Ab (antibody). Spike-f (spike foldon), NC-C (nucleocapsid c-terminal chain).

**Figure 1.**
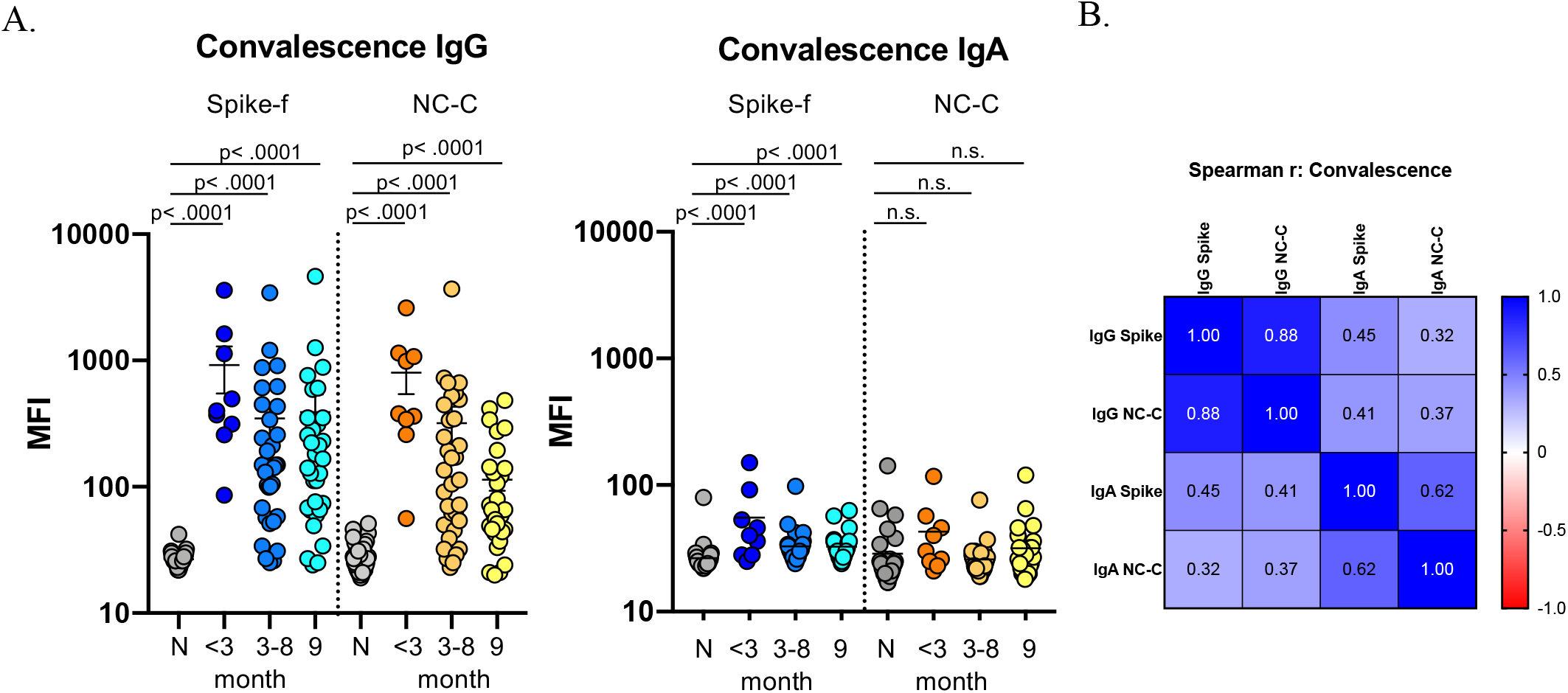
Measurement of IgG and IgA to Spike-f (soluble trimeric form of the spike glycoprotein stabilized in the pre-fusion conformation) and NC-C (nucleocapsid C-terminal fragment) of SARS-CoV2 in saliva of convalescence patients (Cohort 1). (A) Multiplex assay measured signal scores on indicated immunoglobulins to Spike-f and NC-C in cohorts of pre-COVID samples (n=35), and convalescent patient samples at indicated month post infection (n=74). The data is expressed in median fluorescence index (MFI) and plotted using dot plots where each dot is one individual sample. Horizontal bars denote the mean and vertical line represents standard errors. Mann-Whitney U test for significance was performed (B) Spearman correlation analysis with coefficient indicated for respective antibody specificity pairs. n.s = not significant.

Moreover, salivary IgG to Spike-f and NC-C showed to be highly correlated in this cohort (r=0.88, p<0.0001, Spearman correlation test), with concordant serostatus in the majority of samples (**Figure 1b)**. Significant albeit moderate correlations were also seen between IgA to Spike-f and NC-C (r=0.62, p<0.001), and between Spike-f specific IgA and IgG (r=0.45, p<0.001) (Figure 1b).

### Salivary antibody reactivity to SARS-CoV-2 in healthy donors is associated with the recent history of COVID-19 like symptoms

Next, we applied this assay platform to evaluate a second independent cohort - Cohort 2. Participants here were self-reporting symptom-free individuals visiting the University Dental Clinic’s premises of Karolinska Institutet and the Eastman Institute in Stockholm. A total of 147 individuals from May to November 2020 participated in and donated saliva samples. Samples were collected and tested using the same standard operating protocol as for Cohort 1. Shown in **Figure 2a**, and based on antigen-specific cutoffs calculated on 20 negative controls, antibody reactivities to Spike-f and NC-C in this cohort were as following: IgG were detected in 14% to either Spike-f or NC-C, while 11% had detectable IgG to both antigens; for IgA, 10% and 6% of the samples showed reactivity to Spike-f and NC-C respectively, while only 5% showed reactivity to both. Salivary positivity was particularly enriched among participants with self-reported recent history of COVID-19-like symptoms (14 days to 3 months prior to sampling time). Significant reactivities of IgG (p=0.004, and p=0.01) and IgA (p<0.0001, and p=0.044) to either Spike-f or NC-C was found to associate with recent history of symptoms compared to pre-pandemic controls (**Figure 2a**).

**Figure 2.**
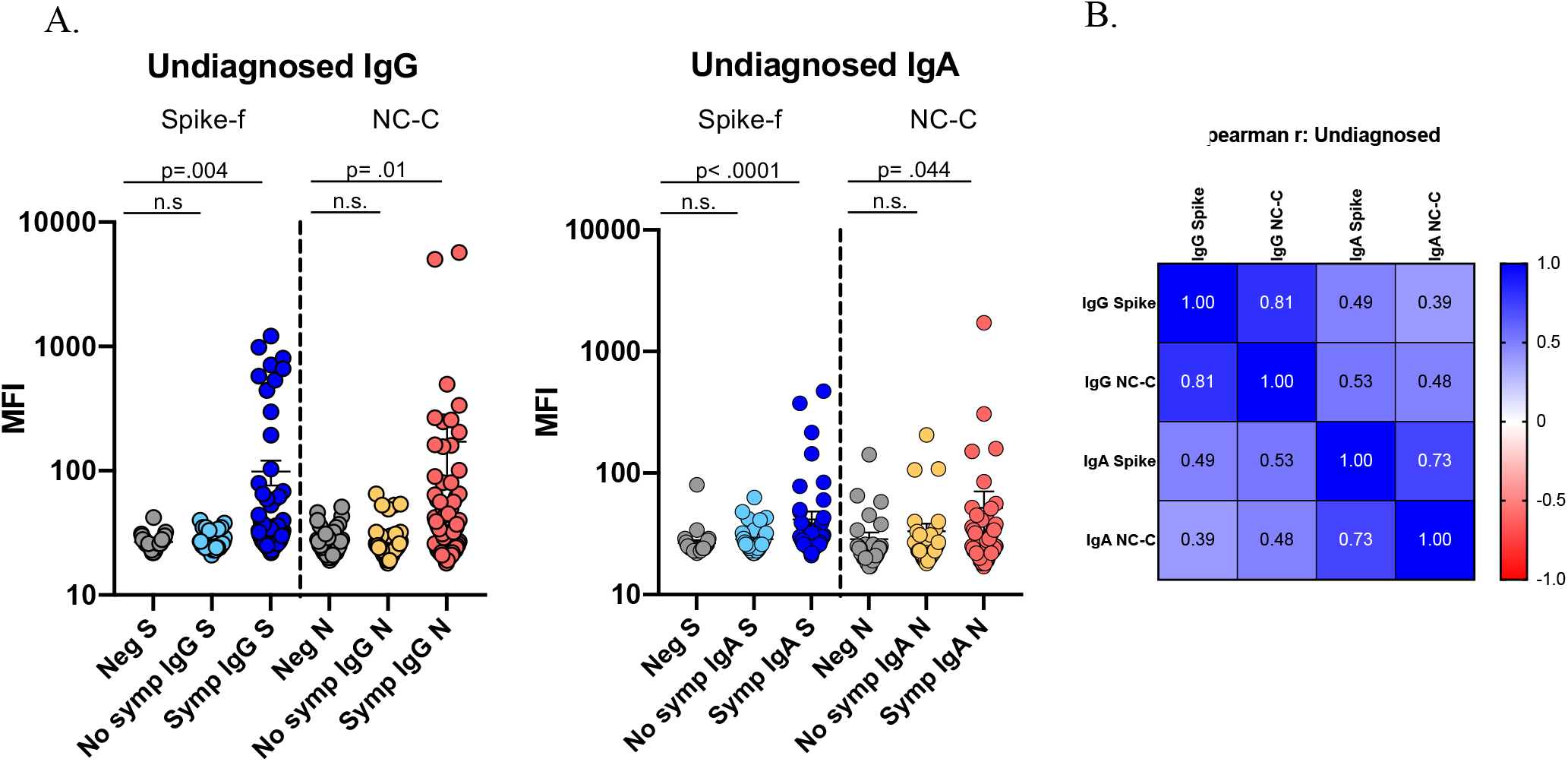
SARS-CoV2 specific IgG and IgA in saliva of undiagnosed study participants (Cohort 2) measured by same method as in Figure 1. Samples were sub-grouped by participant reported COVID-like symptoms the past 14 days - 3 months prior to sampling (Cohort 2). (A) Multiplex assay measured signal scores on indicated immunoglobulins to Spike-f and NC-C in cohorts of pre-COVID samples (n=35), and convalescent patient samples at indicated month post infection (n=146). The data is expressed in median fluorescence index (MFI) and plotted using dot plots where each dot is one individual sample. Horizontal bars denote the mean and vertical line represents standard errors. Mann-Whitney U test for significance was performed (B) Spearman correlation analysis with coefficient indicated for respective antibody specificity pairs. n.s = not significant.

A correlation analysis (**Figure 2b**) gave similar result as for Cohort 1, with highest reported correlation between salivary IgG to Spike-f and NC-C (r=0.81, p<0.0001, Spearman correlation test). Significant albeit moderate correlations were also seen between IgA to Spike-f and NC-C (r=0.73, p<0.001), and IgG and IgA to Spike-f (r=0.49, p<0.001), and Spike-f IgA to NC-C IgG (r=0.53, p<0.001).

### Saliva antibody stability – the influence of inactivation pre-treatment and room temperature

Next the effects of inactivation treatment with 1% Triton X-100 or heat-treatment at 56 C, as well as room temperature storage (identical aliquots left out for indicated time) on the antibody results were determined (**Figure 3)**. Both 1% Triton X-100 and heat treatment showed slight or no change in the cutoff calculated based on the ten included negative controls. A good correlation between treated and non-treated samples was noted **(Figure 3 and S1)**, with a few exceptions of single samples that show a drop in IgG reactivity. Simulation with room temperature storage (22°C) showed a slow decay in IgG signal intensity in positive samples (blue) over time, with the signals of negative samples remain low and stable (grey). Based on these data, inactivation by Triton X-100 or heat treatment treatments seems to have little effect on saliva samples. However, antibody decay variations showed slight IgG signal reduction by each day of room temperature storage.

**Figure 3.**
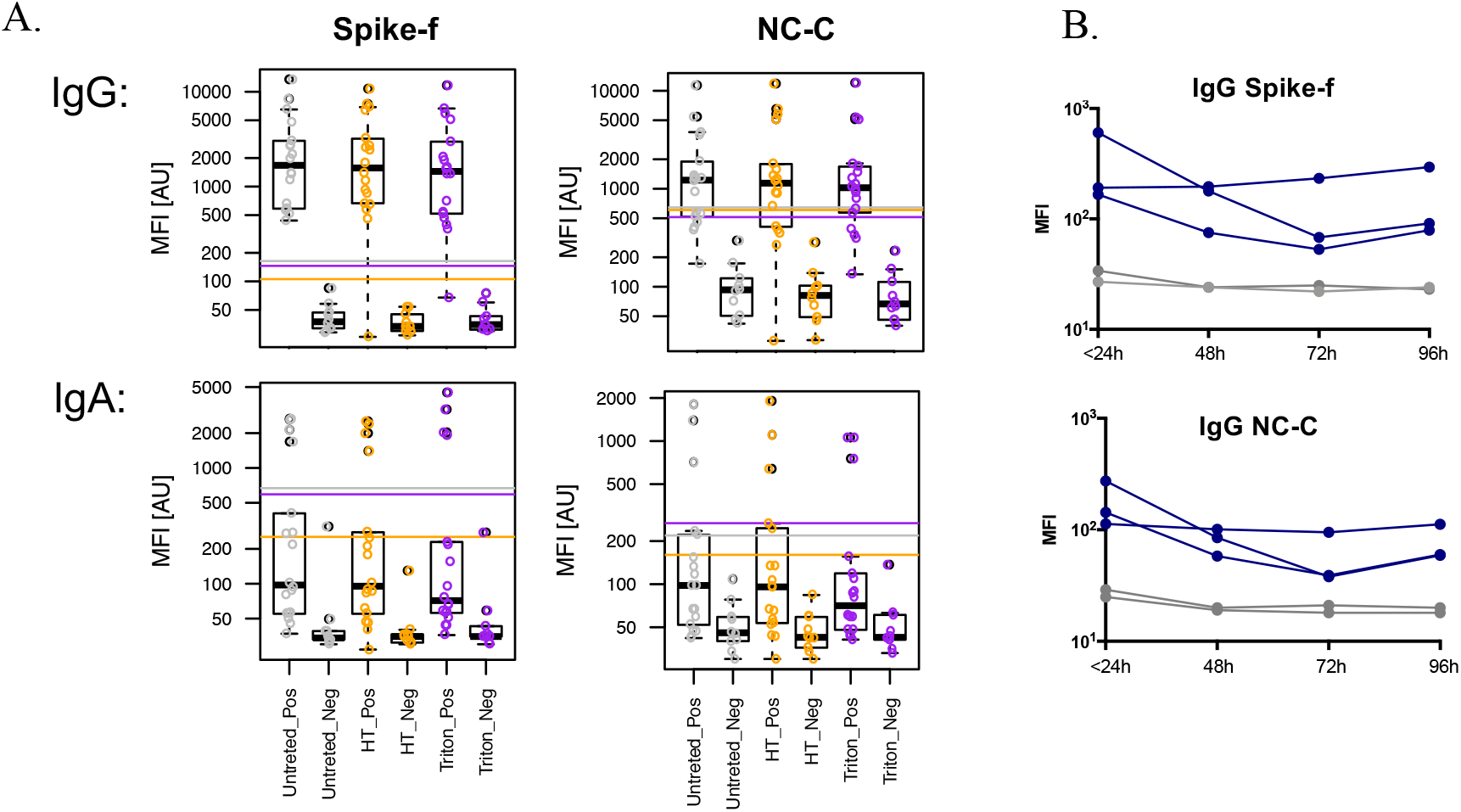
Stability tests of saliva samples subjected to heat (HT), 1% Triton-X-100 (Triton), or left for indicated time in room temperature. (A) SARS-CoV2 specific IgG and IgA reactivities in convalescence saliva samples (Pos) or Pre-pandemic saliva samples (Neg) were tested either untreated (Untreated) or after heat inactivation at 56°C for 30 min (HT), or after Triton-X-100 inactivation (final 1% volume/volume) for 60 min (Triton). Reactivities to Spike-f respective NC-C antigens are shown as box plots with each dot representing one single sample. (B) Convalescence saliva samples (blue) or pre-pandemic saliva samples (grey) were aliquoted and placed in room temperature (22°C) at indicated time points, thereafter snap frozen and tested in same assay run for measurement of SARS-CoV2 specific IgG to spike or nucleocapsid.

## Discussion

Comprehensive antibody testing and the subsequent interventions they generate are essential to monitoring and control SARS-CoV-2 transmission. The present study demonstrates that SARS-CoV-2 specific IgG in saliva after mild COVID-19 can serve as a complementary measure of exposure or immunity to SARS-CoV-2, particularly due to their frequent concurrence with serum IgG responses. Key findings include 1) SARS-CoV-2-specific mucosal salivary antibodies co-exist with the circulating blood antibodies up to 9 months post-natural infection in the majority of participants (88% in saliva vs. 97% in blood); 2) natural infection induces salivary antibodies to recognize both viral spike and nucleocapsid proteins; 3) the response correlates significantly to recent Covid-19-like symptom history in undiagnosed individuals; 4) saliva IgG is relatively stable tolerating both biosafety required temperature and detergent pre-treatment. All together representing a non-invasive approach suitable for population-based immunity surveys. Ideally, if the latter is sampled at home and mailed to the lab, it can help protect vulnerable persons at risk for severe COVID-19 by sparing the need to visit the laboratory units for blood drawls. This is an appealing way to test persons, in pandemic situation, and definitely a complementary test for conventional blood IgG assay. Our data also showed that sample inactivation with either heat treatment or Triton X-100 might be both safe options for testing saliva sample in the lab, causing very little to no variation on the assay performance.

Severe COVID-19 symptoms have been shown to cause strong antibody responses in 99% of convalescence individuals, but published data show also that the antibody responses tend to disappear faster in cases with mild symptoms (6, 9, 16, 19). Possible reason for this is that tests developed earlier during the pandemic were based on detection in samples from severe COVID-19 cases rather than individuals with mild symptoms, hence sensitivity was not optimal (23). Further, many of the early developed tests are using the nucleocapsid as antigen and antibodies targeting this part of the virus has been shown to decline faster (24) as also was detected here. In this study, we deliberately recruited convalescence samples from mild COVID-19 patients, showing that the multiplex antibody platform used here was capable of detecting specific SARS-CoV-2 antibodies in saliva up to 9 months post infection. In the present study, the clinical blood test results that were compared with the saliva reactivities are from certified patient diagnostics (including anti-N pan-Ig ECLIA), showed high performance in detecting late convalescence blood samples. In fact, our result is in line with a recent South Korean group reporting this diagnostic antibody assay is, among several others, effective in detecting SARS-CoV-2 antibodies in blood (90%) of individuals up to 8 months after either asymptomatic infection or reporting mild-symptoms (25). The persistence of salivary IgG to structural viral proteins in the saliva samples after 9 months recovery from mild COVID-19 is intriguing, and possibly explained by a secondary exposure or spill-over from the blood. More studies are therefore warranted to clarify this. In relation to it, the mucosal antibody response is triggered slightly earlier upon infection (10). Information is still limited about the duration and kinetics of mucosal antibodies secreted into the mouth and nose, particularly in this patient group. A sensitive salivary antibody detection assay with the capability to identify infections with various severities would contribute to improving the current understanding of mucosal antibodies to SARS-CoV-2. For instance, such studies may compare low versus high avidity antibodies and their relation to neutralization or disease enhancement (10,26,27).

The notion that antibodies to previously known coronaviruses may block SARS-CoV-2 has raised a concern about whether these antibodies are functional. However, such antibodies are also known to be protective only for about 6 months after the infection, therefore would have disappeared in most cases of SARS-CoV-2 (28,29), Comparing saliva samples using pseudo-neutralizing assay in ACE-2 cross-blocking experiments will therefore be interesting. Other applications for quantitative and qualitative saliva antibody assays include immunity studies to elucidate vaccine-induced mucosal immunity, including the response to antigens representing new virus mutants and vaccine-induced escape mutants. Since mouth and nose are the first port of entry for SARS-CoV-2, sensitive and accurate methods for quantitative measurements of such local mucosal immunity will lead to better means to combat this virus.

One limitation of our study was the relatively small sample size and the predominantly male population. Another weakness is blood samples were not analyzed same way as saliva, and as several diagnostic assays were used only binary data is given. Also, because of the cross-sectional design, we could not obtain baseline or longitudinal saliva samples. Moreover, we could not assess individual possibilities of re-exposure or re-infection. However, it is unlikely that humoral immunity was boosted because in Stockholm, where the study takes place, the period June-Nov 2020 (second-wave) showed an increase in the daily incidence rate of COVID-19 from 30 to 400 cases/100,000 population (30). In conclusion, despite waning immunity concerns, the present study shows how our multiplex bead-based immunoassays can detect antibodies against SARS-CoV-2 in saliva collected at 9 months after infection in the majority of mildly symptomatic persons.

## Data Availability

The datasets generated during the current study are available from the corresponding author on reasonable request.

## Authors contributions

E.P., A.M., P.N., M.S.C. conception and design of the study. H.A., S.B., A.M., A.O., collected the material and performed the experiments. H.A., S.B., C.H. A.M., E.P., M.S.C. analyzed the data. E.P., AM, P.N., and M.S.C. supervised the work. K.L., S.A. G.B. S.H. contributed with material and data interpretations, H.A., S.B., E.P., and M.S.C. wrote the manuscript. All authors reviewed and revised the manuscript critically.

## Acknowledgments

All study participants who took interest in this study.

## Supplementary materials

**Supplementary Table 1.**
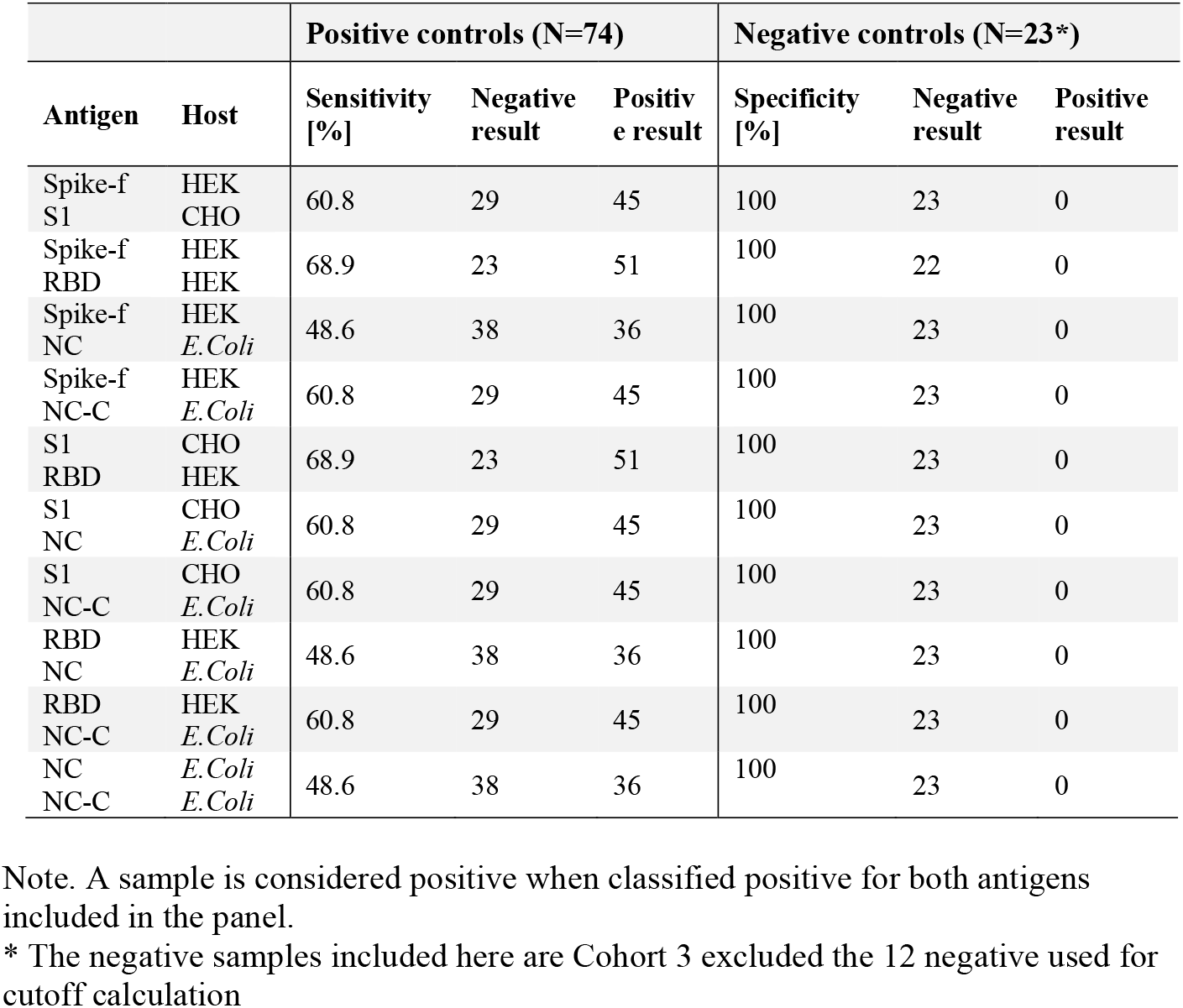
Combinations of 2 antigens. Specificity and sensitivity in detecting SARS-CoV-2 IgG in Cohort 1 samples (1-9 months convalescence).

**Supplementary Table 2.** Combinations of 3 antigens. Specificity and sensitivity in detecting SARS-CoV-2 IgG in Cohort 1 samples (1-9 months convalescence).

|  |  | Positive controls (N=74) |  |  | Negative controls (N=23*) |  |  |
| --- | --- | --- | --- | --- | --- | --- | --- |
| Antigen | Host | Sensitivity [%] | Negative result | Positive result | Specificity [%] | Negative result | Positive result |
| Spike-f | HEK | 70.3 | 22 | 52 | 100 | 23 | 0 |
| S1 | CHO |  |  |  |  |  |  |
| RBD | HEK |  |  |  |  |  |  |
| Spike-f | HEK | 62.2 | 28 | 46 | 100 | 23 | 0 |
| S1 | CHO |  |  |  |  |  |  |
| NC | *E.Coli* |  |  |  |  |  |  |
| Spike-f | HEK | 64.9 | 26 | 48 | 100 | 23 | 0 |
| S1 | CHO |  |  |  |  |  |  |
| NC-C | *E.Coli* |  |  |  |  |  |  |
| S1 | CHO | 62.2 | 28 | 46 | 100 | 23 | 0 |
| RBD | HEK |  |  |  |  |  |  |
| NC | *E.Coli* |  |  |  |  |  |  |
| RBD | HEK | 58.1 | 31 | 43 | 100 | 23 | 0 |
| NC | *E.Coli* |  |  |  |  |  |  |
| NC-C | *E.Coli* |  |  |  |  |  |  |
| S1 | CHO | 58.1 | 31 | 43 | 100 | 23 | 0 |
| NC | *E.Coli* |  |  |  |  |  |  |
| NC-C | *E.Coli* |  |  |  |  |  |  |
| Spike-f | HEK | 60.8 | 29 | 45 | 100 | 23 | 0 |
| NC | *E.Coli* |  |  |  |  |  |  |
| NC-C | *E.Coli* |  |  |  |  |  |  |
| Spike-f | HEK | 70.3 | 22 | 52 | 95.7 | 22 | 1 |
| RBD | HEK |  |  |  |  |  |  |
| NC-C | *E.Coli* |  |  |  |  |  |  |
| S1 | CHO | 59.5 | 30 | 44 | 100 | 23 | 0 |
| RBD | HEK |  |  |  |  |  |  |
| NC-C | *E.Coli* |  |  |  |  |  |  |
| Spike-f | HEK | 67.6 | 24 | 50 | 95.7 | 22 | 1 |
| RBD | HEK |  |  |  |  |  |  |
| NC | *E.Coli* |  |  |  |  |  |  |

Note. A sample is considered positive when classified positive for two out of three antigens included in the panel.
\* The negative samples included here are Cohort 3 excluded the 12 negative used for cutoff calculation

**Supplementary Table 3.** Single antigen specificity and sensitivity in detecting SARS-CoV-2 IgA in Cohort 1 samples (1-9 months convalescence).

|  |  | Positive controls (N=74) |  |  | Negative controls (N=23*) |  |  |
| --- | --- | --- | --- | --- | --- | --- | --- |
| Antigen | Host | Sensitivity [%] | Negative result | Positive result | Specificity [%] | Negative result | Positive result |
| Spike-f* | HEK | 16.2 | 62 | 12 | 95.7 | 22 | 1 |
| S1 | CHO | 5.4 | 70 | 4 | 91.3 | 21 | 2 |
| RBD | HEK | 6.8 | 69 | 5 | 91.3 | 21 | 2 |
| NC | *E. coli* | 6.8 | 69 | 5 | 87.0 | 20 | 3 |
| NC-C* | *E. coli* | 5.4 | 70 | 4 | 91.3 | 21 | 2 |

\* The negative samples included here are Cohort 3 excluded the 12 negative used for cutoff calculation

**Figure S1.**
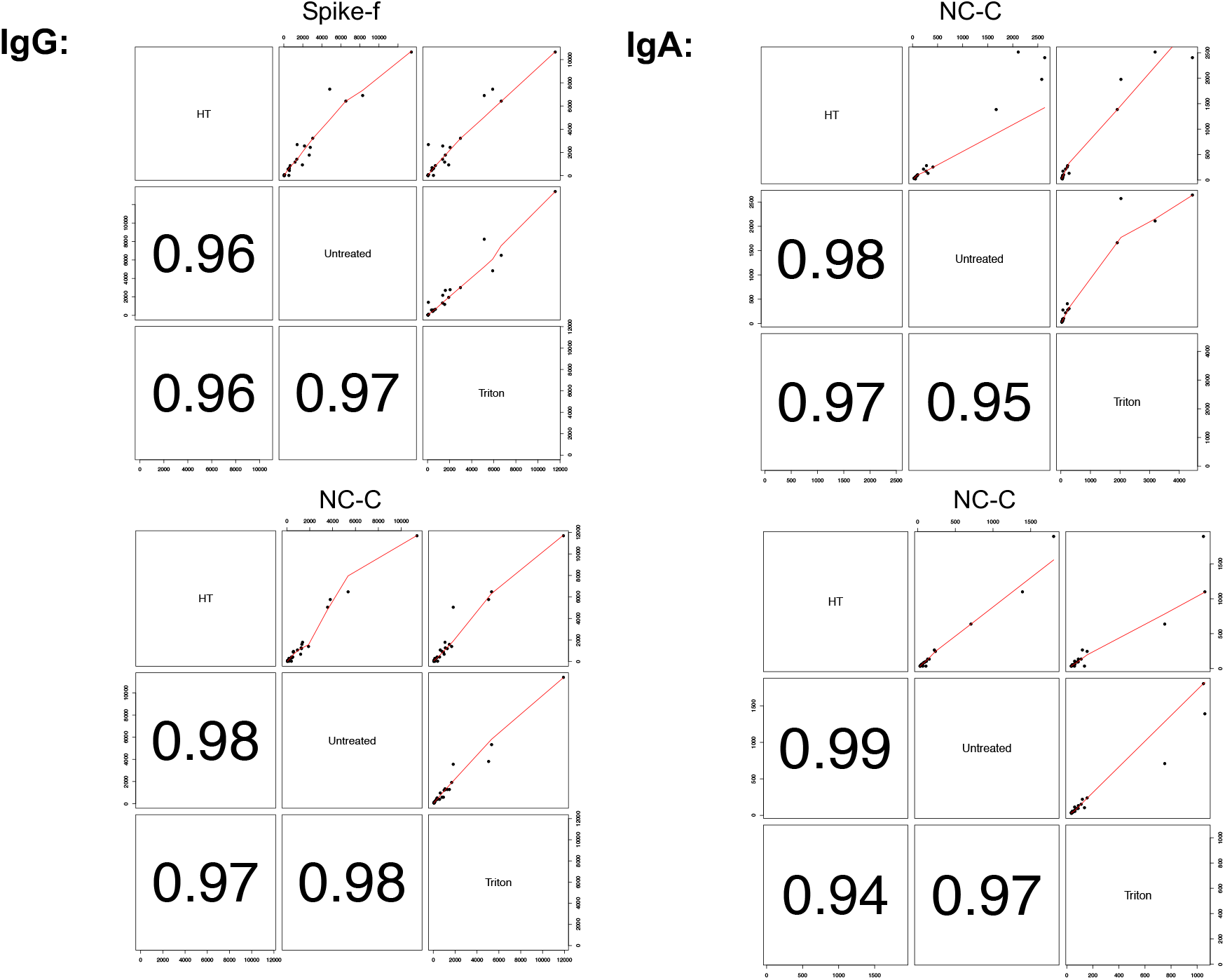
Correlation analysis comparing inactivation by heat, Triton-X-100, and untreated conditions as shown in Figure 3A.

